# Association between DNA Methylation Levels in Brain Tissue and Late-Life Depression in Community-Based Participants

**DOI:** 10.1101/2020.04.21.20074021

**Authors:** Anke Hüls, Chloe Robins, Karen N. Conneely, Philip L. De Jager, David A. Bennett, Michael P. Epstein, Thomas S. Wingo, Aliza P. Wingo

## Abstract

**Objective:** Major depressive disorder (MDD) arises from a combination of genetic and environmental risk factors and DNA methylation is one of the molecular mechanisms through which these factors can manifest. However, little is known about the epigenetic signature of MDD in brain tissue. This study aimed to investigate associations between brain tissue-based DNA methylation and late-life MDD.

**Methods:** We performed a brain epigenome-wide association study (EWAS) of late-life MDD in 608 participants from the Religious Order Study and the Rush Memory and Aging Project (ROS/MAP) using DNA methylation profiles of the dorsal lateral prefrontal cortex (dPFC) generated using the Illumina HumanMethylation450 Beadchip array. We also conducted an EWAS of MDD in each sex separately.

**Results:** We found epigenome-wide significant associations between brain-tissue-based DNA methylation and late-life MDD. The most significant and robust association was found with altered methylation levels in the *YOD1* locus (cg25594636, p-value=2.55 × 10^−11^; cg03899372, p-value=3.12 × 10^−09^; cg12796440, p-value=1.51 × 10^−08^, cg23982678, p-value=7.94 × 10^−08^). Analysis of differentially methylated regions (DMR, p-value=5.06 × 10^−10^) further confirmed this locus. Other significant loci include *UGT8* (cg18921206, p-value=1.75 × 10^−08^), *FNDC3B* (cg20367479, p-value=4.97 × 10^−08^) and *SLIT2* (cg10946669, p-value=8.01 × 10^−08^). Notably, brain-tissue based methylation levels were strongly associated with late-life MDD in men more than in women.

**Conclusions:** We identified altered methylation in the *YOD1, UGT8, FNDC3B* and *SLIT2* loci as new epigenetic factors associated with late-life MDD. Furthermore, our study highlights the sex-specific molecular heterogeneity of MDD.

## Introduction

Major depressive disorder (MDD) severely limits psychosocial functioning, diminishes quality of life and is a leading cause of disability worldwide ^1^. The 12-month prevalence of MDD is approximately 6% ^2^ and similar when comparing high-income countries with low-income and middle-income countries, indicating that MDD is neither a simple consequence of modern day lifestyle in developed countries, nor poverty ^3,4^. Furthermore, although social and cultural factors such as socioeconomic status can have a role in major depression, genomic and other underlying biological factors ultimately drive the occurrence of this condition ^5^. Twin studies have provided heritability estimates of the MDD of approximately 30–40% ^6^. One of the molecular mechanisms through which environmental and genetic factors can modulate a disease outcome is epigenetics, with DNA methylation being one of the most studied modifications of the genome.

Recent epigenome-wide association studies (EWAS) showed an association of whole-blood DNA methylation levels with depressive symptoms ^7,8^ as well as MDD ^9–11^, but little is known about brain epigenetic markers of depression or MDD. The EWAS of depressive symptoms were both conducted in late middle-aged and elderly people from the general population (mean age 70 years ^8^ and 65 years ^7^), an age group with an increased risk of developing dementia ^12^. However, both studies could not determine if their findings were confounded by dementia, which is known to be highly associated with late-life depression ^13^. On the other hand, recent EWAS of MDD were performed in younger participants (mean age 42 years) ^9–11^ and it is unclear if their findings can be generalized across age groups. Furthermore, most previous EWAS on depression were limited due to measuring DNA methylation changes in blood ^7–10^. Two recent studies conducted EWAS of MDD in 206 postmortem brain samples, but the MDD diagnosis was based on information obtained from a family member and there was no information on dementia ^9,11^. Thus, there is need to understand the epigenetic changes in the human brain that are associated with late-life MDD and to determine if these changes are independent of dementia.

In this study, we investigated associations between both brain tissue-based individual CpGs as well as regions of differential methylation and late-life MDD in 608 participants from the Religious Order Study and Rush Memory and Aging Project (ROS/MAP) cohorts. To reduce the risk of confounding by cognitive status, we excluded participants with a diagnosis of dementia at the time of MDD assessment and adjusted for cognitive status at the last follow-up visit (closest to methylation assessment) in our analyses. Furthermore, we performed a stratified analysis for men and women to investigate the sex-specific methylation patterns of MDD.

## Methods

### Study design and study population

The study population included deceased subjects from two large, prospectively followed cohorts recruited by investigators at Rush Alzheimer’s Disease Center in Chicago, IL: The Religious Orders Study (ROS) and the Rush Memory and Aging Project (MAP) ^14,15^. Participants provided informed consent, an Anatomic Gift Act for organ donation, and a repository consent to allow their data to be repurposed. Both studies were approved by an Institutional Review Board of Rush University Medical Center. To be included in the present study, participants must have been assessed for major depressive disorder and have available genotype data and methylation profiles derived from the dorsolateral prefrontal cortex. Furthermore, we excluded participants with a diagnosis of dementia at the time of MDD assessment (at baseline evaluation). As in previous publications, the ROS and MAP data were analyzed jointly since much of the phenotypic data collected are identical at the item level in both studies and collected by the same investigative team ^14,16^.

### DNA methylation

DNA methylation was measured from the dorsolateral prefrontal cortex (dPFC; Broadman area 46) as previously described in 737 ROS/MAP participant samples^14^. DNA was extracted from cortically dissected sections of dPFC and DNA methylation was measured using the Illumina HumanMethylation450 Beadchip array. Initial data processing, including color channel normalization and background removal, was performed using the Illumina GenomeStudio software. The raw IDAT files were obtained from Synapse (www.synapse.org;Synapse ID: syn7357283) and the following probes were removed: 1) probes with a detection p-value > 0.01 in any sample, 2) probes annotated to the X and Y chromosomes by Illumina, 3) probes that cross-hybridize with other probes due to sequence similarity, 3) non-CpG site probes, and 4) probes that overlap with common SNPs. After this filtering, the remaining CpG sites were normalized using the BMIQ algorithm in Watermelon R package ^17^, and the ComBat function from the sva R package was used to adjust for batch effects ^18^. After quality control 408,689 discrete CpG dinucleotides in 608 subjects were used for analysis.

### Genotype data

Genotyping data was generated using two microarrays, Affymetrix GeneChip 6.0 (Affymetrix, Inc, Santa Clara, CA, USA) and Illumina HumanOmniExpress (Illumina, Inc, San Diego, CA, USA) as described previously ^19^. Genotyping was imputed to the 1000 Genome Project Phase 3 using the Michigan Imputation Server ^20^, and the following filtering criteria were applied minor allele frequency (MAF) > 5%, Hardy-Weinberg p-value >10^−5^ and genotype imputation R^2^ > 0.3. Principal components were estimated using common (MAF>0.05) unlinked (R^2^<0.1) autosomal markers by EIGENSTRAT^21^.

### Diagnosis of major depressive disorder

A clinical diagnosis of current major depressive disorder was rendered by an examining clinician. The diagnosis was based on clinical interview using the criteria of the Diagnostic and Statistical Manual of Mental Disorders, 3rd Edition, Revised (DSM-III-R) ^22^. The MDD diagnosis included present versus not present. In this study, we focused on the diagnosis of MDD at the baseline assessment to reduce the risk that our findings are confounded by dementia.

### Clinical diagnosis of cognitive status

A clinical diagnosis of dementia status was rendered based on a three-stage process including computer scoring of cognitive tests, clinical judgment by a neuropsychologist, and diagnostic classification by a clinician. All participants undergo a uniform, structured, clinical evaluation including a battery of 21 cognitive tests of which 19 are in common. These tests were scored by computer using a decision tree designed to mimic clinical judgment and a rating of severity of impairment was given for 5 cognitive domains. A neuropsychologist, blinded to participant demographics, reviews the impairment ratings and other clinical information and renders a clinical judgment regarding the presence of impairment and dementia. A clinician (neurologist, geriatrician, neuropsychologist, or geriatric nurse practitioner) then reviews all available data and examines the participant and renders a final diagnostic classification. Clinical diagnosis of dementia and clinical Alzheimer’s disease (AD) are based on criteria of the joint working group of the National Institute of Neurological and Communicative Disorders and Stroke and the Alzheimer’s Disease and Related Disorders Association (NINCDS/ADRDA). The diagnosis of AD requires evidence of a meaningful decline in cognitive function relative to a previous level of performance with impairment in memory and at least one other area of cognition. Diagnosis of mild cognitive impairment (MCI) is rendered for persons who are judged to have cognitive impairment by the neuropsychologist but are judged to not meet criteria for dementia by the clinician. Persons without dementia or mild cognitive impairment (MCI) are categorized as having no cognitive impairment (NCI).

### Statistical analysis

For the brain epigenome-wide association study (EWAS) of MDD, we ran a multivariate robust linear regression model with empirical Bayes from the R package limma (version 3.40.6) ^23^ using clinical diagnosis of MDD at baseline as the independent variable and each CpG methylation as a dependent variable, adjusting for age at death, sex, postmortem interval (PMI), proportion of neurons and the first three genetic principal components. The effect estimates from the adjusted models (**Δ** beta) refer to the difference in mean DNA methylation beta values between groups (with and without MDD). We applied a Bonferroni threshold to correct for multiple testing based on the number of tested CpG sites (threshold: 0.05/408,689 = 1.22 × 10^−07^). Fine-mapping of our epigenome-wide associations was done with coMET ^24^, which is a visualization tool of EWAS results with functional genomic annotations and estimation of co-methylation patterns. We conducted the following sensitivity analyses: 1) We included the cognitive status at the last follow-up visit (closest to methylation assessment) as a covariate to investigate if our findings were confounded by dementia, 2) We confirmed our associations using linear regression with p-values obtained from normal theory (lm() function in R) as well as from a permutation test, 3) We corrected the p-values for inflation and bias using a Bayesian method for estimation of the empirical null distribution as implemented in the R/Bioconductor package *bacon* ^25^, and 4) We adjusted our association models for a polygenic risk score for MDD (calculated with PRSice ^26^ and UK Biobank summary statistics from ^27^ with a p-value < 0.05) to test if our EWAS findings were independent of genetic risk for MDD.

CpG sites that reach epigenome-wide significance were replicated using the summary statistics from a cell type-specific EWAS of MDD, which is based on methylation enrichment-based sequencing data from three collections of human postmortem brain (n = 206) ^11^. This replication was used to validate our findings and to provide mechanistic insights about the most relevant cell types for our associations.

Differentially methylated regions (DMRs) in MDD were identified using DMRcate, that identifies DMRs from tunable kernel smoothing process of association signals ^28^. Input files were our single-CpG EWAS results on MDD including regression coefficients, standard deviations and uncorrected p-values. DMRs were defined based on the following criteria: a) a DMR should contain more than one probe; b) regional information can be combined from probes within 1,000 bp; c) the region showed FDR corrected p-value < 0.05.

To identify plausible pathways associated with MDD, we performed an over-representation analysis based on the 1,000 CpGs with the lowest p-values for the association with MDD. We used the R Bioconductor package missMethyl (version 1.18.0 gometh function), which performs one-sided hypergeometric tests taking into account and correcting for any bias derived from the use of differing numbers of probes per gene interrogated by the array ^29^.

## Results

### Description of Study Participants

There were 608 ROS/MAP participants included in this study with an average age at baseline visit of 81 years and an average age of death of 86 years (Table 1). Sixty-four percent of the participants were female. At baseline, 5% of the participants were diagnosed with MDD, which is consistent with the twelve-month prevalence rate of MDD in the general population^2^.

**Table 1.** Study characteristics.

|  | All | Male | Female |
| --- | --- | --- | --- |
| N | 608 | 220 | 388 |
| Age at baseline visit | 80.55 ± 6.51 | 78.76 ± 6.72 | 81.57 ± 6.17 |
| Age at death, mean ± sd | 86.31 ± 4.73 | 84.94 ± 5.41 | 87.09 ± 4.11 |
| Female, n (%) | 388 (63.82%) | 0 (0.00%) | 388 (100.00%) |
| Post mortem interval (PMI), mean ± sd | 7.55 ± 6.01 | 7.80 ± 7.50 | 7.41 ± 4.98 |
| Proportion of neurons, mean ± sd | 0.45 ± 0.06 | 0.44 ± 0.06 | 0.45 ± 0.06 |
| Clinical diagnosis of cognitive status at baseline visit |  |  |  |
| No cognitive impairment, n (%) | 396 (65.13%) | 142 (64.55%) | 254 (65.46%) |
| Mild cognitive impairment (MCI), n (%) | 212 (34.87%) | 78 (35.45%) | 134 (34.54%) |
| Alzheimer's disease dementia (AD)<sup>#</sup>, n (%) | 0 (0.00%) | 0 (0.00%) | 0 (0.00%) |
| Other dementia<sup>#</sup>, n (%) | 0 (0.00%) | 0 (0.00%) | 0 (0.00%) |
| Clinical diagnosis of cognitive status at last follow-up visit |  |  |  |
| No cognitive impairment, n (%) | 232 (38.16%) | 93 (42.27%) | 139 (35.82%) |
| Mild cognitive impairment (MCI), n (%) | 177 (29.11%) | 66 (30.00%) | 111 (28.61%) |
| Alzheimer's disease dementia (AD), n (%) | 184 (30.26%) | 53 (24.09%) | 131 (33.76%) |
| Other dementia<sup>#</sup>, n (%) | 8 (1.32%) | 5 (2.27%) | 3 (0.77%) |
| Clinical diagnosis of MDD at baseline visit | 30 (4.93%) | 9 (4.09%) | 21 (5.41%) |

<sup>#</sup>: Participants with a clinical diagnosis of dementia at baseline were excluded from the analysis sample.

Women showed a slightly higher prevalence of MDD than men (5.4% versus 4.1%, difference not significant).

### Differentially Methylated CpG Sites in Brain Tissue are Associated with Late-Life

*Depression* Differentially methylated CpG sites in the *YOD1* (cg25594636, p-value = 2.55 × 10^−11^; cg03899372, p-value=3.12 × 10^−09^; cg12796440, p-value = 1.51 × 10^−08^, cg23982678, p-value = 7.94 × 10^−08^), *FSTL5* (cg21794994, p-value = 1.46 × 10^−09^), *UGT8* (cg18921206, p-value = 1.51 × 10^−08^), *FNDC3B* (cg20367479, p-value = 4.97 × 10^−08^) and *SLIT2* (cg10946669, p-value = 7.94 × 10^−08^) loci were associated with MDD at the epigenome-wide significance level (Bonferroni-adjustment) after adjusting for sex, PMI, proportion of neurons, first three genetic principal components, and age at death (Table 2, Figure 1 A). These associations were robust to additional adjustment for dementia diagnosis assessed at the last follow-up visit (Table 2). Overall, four CpG sites in *YOD1* were significantly associated with late-life MDD (Table 2) and these were all located in the same CpG island, but only moderately correlated with each other (Figure 2). This CpG island is located in an exon of *YOD1* and in an intron of *PFKFB2*. The distribution of the DNA methylation beta values of the four most significant CpG sites in the *YOD1* locus stratified by MDD diagnosis is shown in Figure S1. The significant associations were confirmed in sensitivity analyses using linear regression models, permutation tests (Table S1) as well as correcting p-values for potential inflation and bias (Figure S2). Furthermore, associations were robust to additional adjustment for a polygenic risk score for MDD, which shows that our EWAS findings were independent of a genetic risk for MDD (Table S2).

**Table 2.** Significant associations between DNA methylation and MDD.

| cpg | chr | position | Nearest Gene | Main model<sup>#</sup> |  | Additionally adjusted for dementia at last follow-up |  |
| --- | --- | --- | --- | --- | --- | --- | --- |
|  |  |  |  | Δ beta | p-value | Δ beta | p-value |
| cg25594636 | 1 | 207224388 | *YOD1* | 0.013 | 2.55E-11 | 0.013 | 2.98E-11 |
| cg03899372 | 1 | 207224102 | *YOD1* | 0.020 | 3.12E-09 | 0.020 | 3.76E-09 |
| cg12796440 | 1 | 207224331 | *YOD1* | 0.022 | 1.51E-08 | 0.022 | 1.34E-08 |
| cg18921206 | 4 | 115320920 | *UGT8* | -0.067 | 1.75E-08 | -0.068 | 9.45E-09 |
| cg20367479 | 3 | 171873675 | *FNDC3B* | -0.032 | 4.97E-08 | -0.032 | 3.89E-08 |
| cg23982678 | 1 | 207224227 | *YOD1* | 0.021 | 7.94E-08 | 0.021 | 7.91E-08 |
| cg10946669 | 4 | 20253130 | *SLIT2* | 0.013 | 8.01E-08 | 0.013 | 6.65E-08 |

Bonferroni threshold: $1.22 \times 10^{-07}$ .
<sup>#</sup>Adjusted for age at death, sex, PMI, neuron subtype proportion and the first three principal components from the genotype data.
Δ beta: This coefficient represents the mean difference of DNA methylation beta values between participants with and without MDD. Negative coefficients refer to smaller mean DNA methylation beta values in participants with MDD and positive coefficients refer to larger mean DNA methylation beta values in participants with MDD.

**Figure 1.**
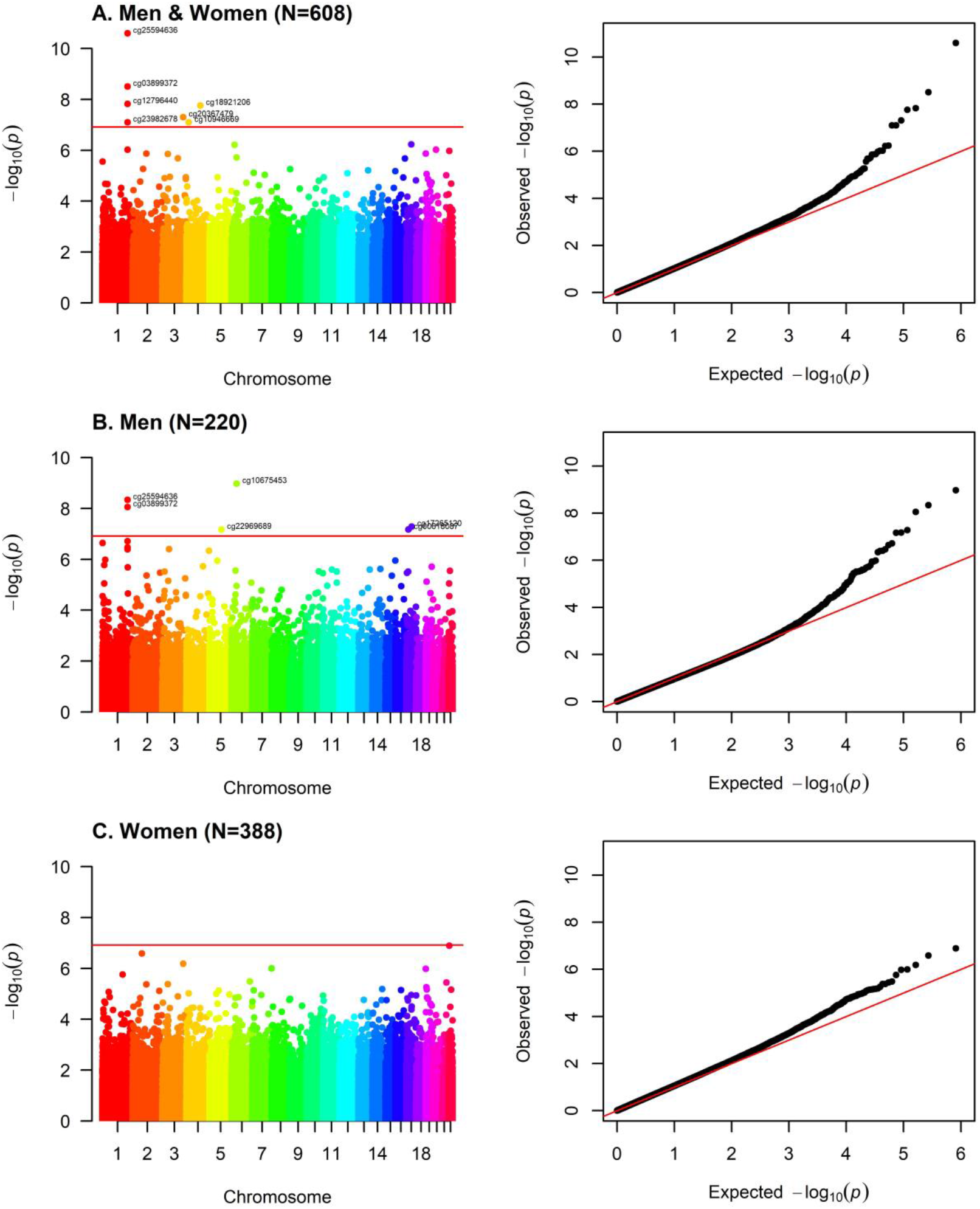
Manhattan and QQ-Plots EWAS on clinical diagnosis of MDD. Adjusted for age at death, sex, PMI, neuron proportion and the first three principal components from the genotype data. Bonferroni threshold: 1.22 × 10^−07^.

**Figure 2.**
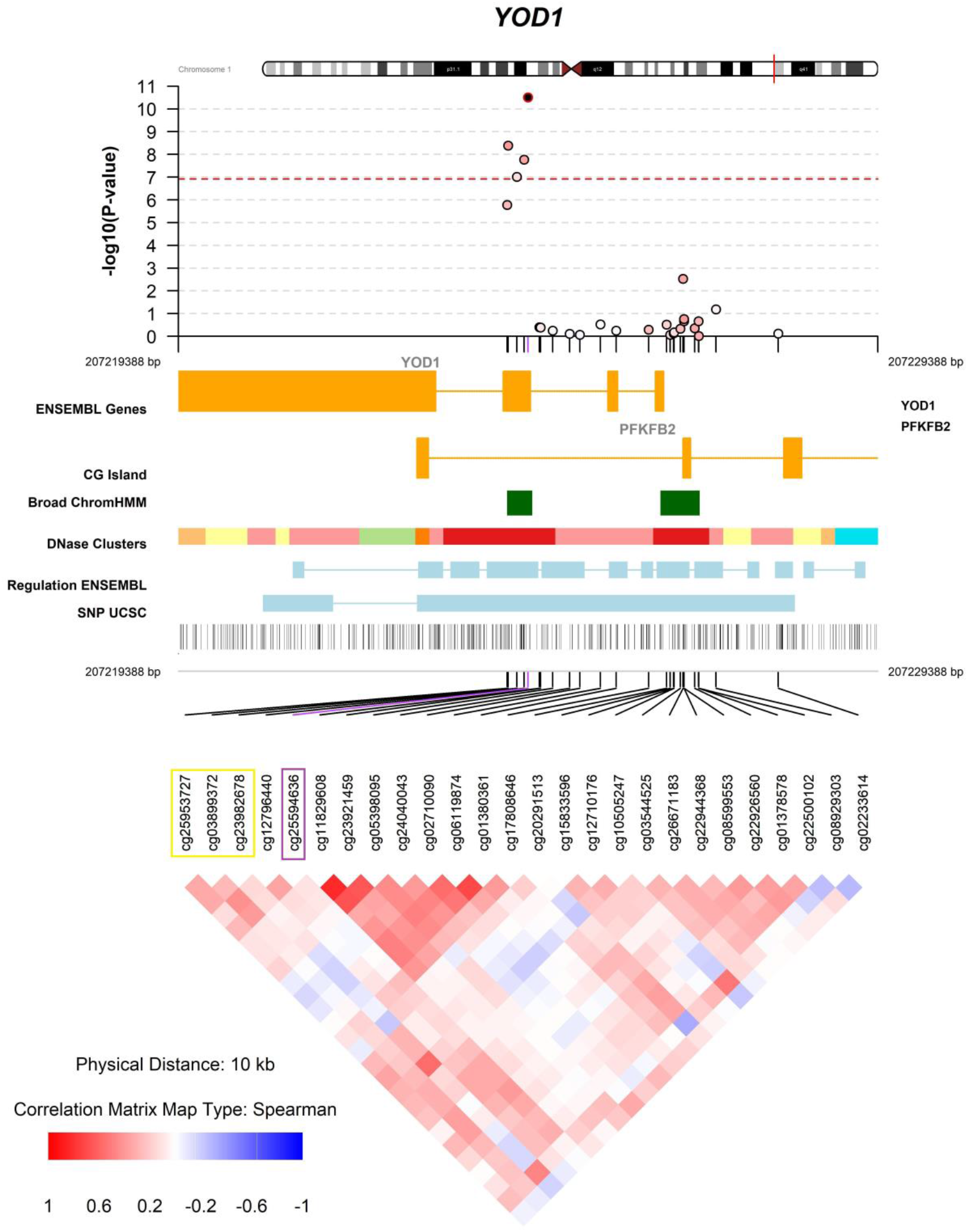
Fine mapping of the association between DNA methylation in *YOD1* and MDD. EWAS results of the association between CpG sites and MDD adjusted for age at death, sex, PMI, neuron proportions and the first 3 principal components from the genotype data. The most significant CpG site (cg25594636) is marked in purple. The three CpG sites marked in yellow belong to a DMR (p-value = 5.06 × 10^−10^, Table S4). The y-axis indicates the strength of association in terms of negative logarithm of the association P value. Each circle represents a CpG site. Red dashed line within the graph indicates the genome-wide significance threshold (Bonferroni threshold: 1.22 × 10^−07^). The regulatory information and correlation matrix of other CpG sites in the region with the top hit are shown below the x-axis. Color intensity marks the strength of the correlation and color indicates the direction of the correlation.

Only two of the seven CpG sites that were significantly associated with MDD in ROS/MAP were included in the cell type-specific EWAS published in ^11^ (cg18921206 and cg20367479, Table S3). Of these, only cg20367479 was nominally significant for bulk brain (p-value = 0.022) and the effects were not robust across different cell types in the replication cohort ^11^ and not in the same direction as in ROS/MAP.

### Analyses of Differentially Methylated Regions

We identified one significant DMR from our EWAS results on late-life MDD that is located in the *YOD1 / PFKFB2* locus (Figure 2, Table S4, minimum FDR p-value for the region = 5.06 × 10^−10^), which is not surprising given the differential CpG site analysis. This DMR includes three CpG sites that are located downstream of the most significant CpG site from our EWAS on late-life MDD (cg25594636, Table 2).

### Associations are Stronger in Men than in Women

Interestingly, we found more methylation sites associated with MDD in men than in women (Figure 1, Table 3), although the sample size was much smaller in men (N=220 men vs. N=388 women). Differentially methylated CpG sites in *YOD1* were more strongly associated with late-life MDD in men than in women (e.g. for cg03899372, men: beta = 0.041, p-value = 8.80 × 10^−09^; women: beta = 0.010, p-value = 0.0024; p-value sex interaction = 4.51 × 10^−06^; Table 3). Methylation in *PRICKLE4* (p-value sex interaction = 1.26 × 10^−09^), *GFAP* (p-value sex interaction = 6.88 × 10^−05^), *RP11-1E3*.*1* (p-value sex interaction = 1.11 × 10^−07^) and *UBB* (p-value sex interaction = 1.54 × 10^−11^) was only associated with MDD in men, but not in women or in both men and women (Table 3).

**Table 3.**
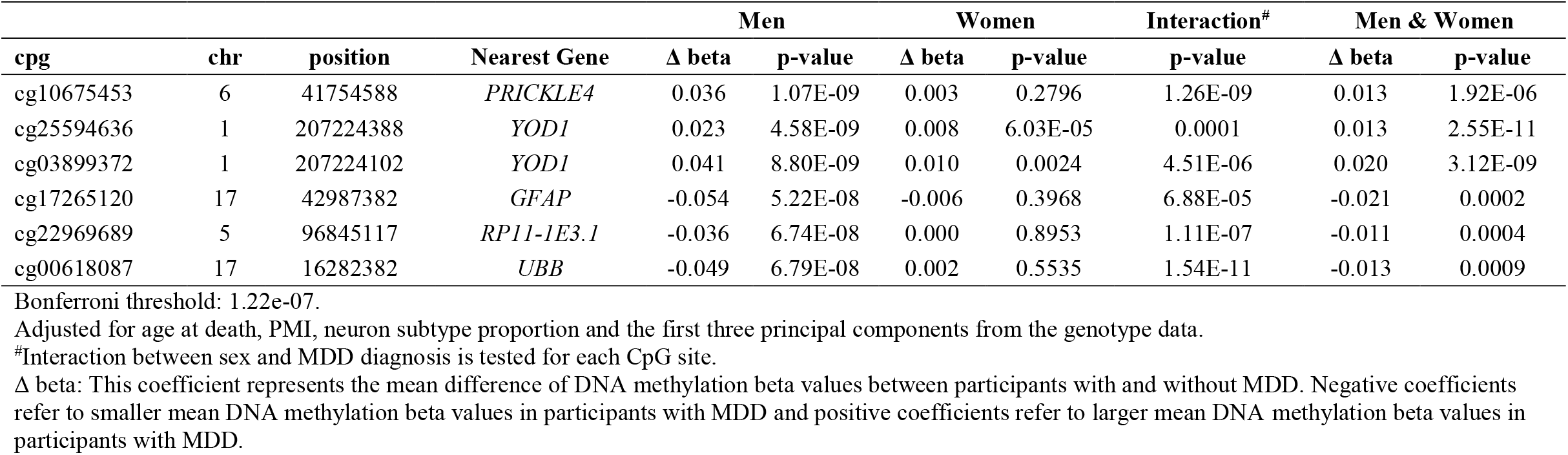
Significant associations between DNA methylation and MDD in male. Associations in men, women and all participants are ordered by the p-values from the analysis of male participants

| cpg | chr | position | Nearest Gene | Men |  | Women |  | Interaction<sup>#</sup> | Men & Women |  |
| --- | --- | --- | --- | --- | --- | --- | --- | --- | --- | --- |
| | | | | $\Delta$ beta | p-value | $\Delta$ beta | p-value | p-value | $\Delta$ beta | p-value |
| cg10675453 | 6 | 41754588 | *PRICKLE4* | 0.036 | 1.07E-09 | 0.003 | 0.2796 | 1.26E-09 | 0.013 | 1.92E-06 |
| cg25594636 | 1 | 207224388 | *YOD1* | 0.023 | 4.58E-09 | 0.008 | 6.03E-05 | 0.0001 | 0.013 | 2.55E-11 |
| cg03899372 | 1 | 207224102 | *YOD1* | 0.041 | 8.80E-09 | 0.010 | 0.0024 | 4.51E-06 | 0.020 | 3.12E-09 |
| cg17265120 | 17 | 42987382 | *GFAP* | -0.054 | 5.22E-08 | -0.006 | 0.3968 | 6.88E-05 | -0.021 | 0.0002 |
| cg22969689 | 5 | 96845117 | *RP11-1E3.1* | -0.036 | 6.74E-08 | 0.000 | 0.8953 | 1.11E-07 | -0.011 | 0.0004 |
| cg00618087 | 17 | 16282382 | *UBB* | -0.049 | 6.79E-08 | 0.002 | 0.5535 | 1.54E-11 | -0.013 | 0.0009 |

Bonferroni threshold: 1.22e-07.
Adjusted for age at death, PMI, neuron subtype proportion and the first three principal components from the genotype data.
<sup>#</sup>Interaction between sex and MDD diagnosis is tested for each CpG site.
$\Delta$ beta: This coefficient represents the mean difference of DNA methylation beta values between participants with and without MDD. Negative coefficients refer to smaller mean DNA methylation beta values in participants with MDD and positive coefficients refer to larger mean DNA methylation beta values in participants with MDD.

### Pathway Analysis

No significantly enriched pathway was found among the 1,000 most significant CpG sites from the EWAS of late-life MDD (Table S5). The smallest p-value (6 × 10^−5^) was reached for calmodulin-dependent protein phosphatase activity (GO:0033192). The genes that belong to this pathway are *PPM1A* (chr12), *PPM1F* (chr16), *PPP3CA* (chr3), *PPP3CB* (chr14) and *PPP3CC* (chr14) (Table S6).

## Discussion

In this study, we found epigenome-wide significant associations between brain-tissue-based DNA methylation and late-life MDD in more than 600 participants from the ROS/MAP cohorts. The most significant and robust association was found with altered methylation levels in the *YOD1 / PFKFB2* loci. This association was not confounded by dementia or a genetic risk for MDD and significant in both the single site and region-based analysis. Interestingly, brain-tissue based methylation levels were stronger associated with late-life MDD in men than in women.

The most significant CpG sites were found in a region covering an exon of *YOD1* and an intron of *PFKFB2. YOD1* is a highly conserved deubiquitinase similar to yeast *OTU1* ^30^ that is associated with regulation of the endoplasmic reticulum (ER)-associated degradation to maintain the proper folded state of proteins ^31^. Additionally, *YOD1* is a negative regulator of TRAF6/p62-triggered IL-1 signaling ^32^ and IL-1 plays an important role in the regulation of inflammatory responses as well as in depression ^33–35^. Together, these suggest that *YOD1* is associated with depression perhaps via influencing the inflammatory responses. Lastly, previous studies suggest that *YOD1* contributes to pathogenesis of neurodegenerative disease like Huntington disease and Parkinson’s disease ^30^. *PFKFB2* has been studied in the context of brain tumors ^36,37^, but there is no evidence for an association with neuropsychological disease. Therefore, we hypothesize that the CpG sites we found to be associated with late-life MDD are most likely linked to *YOD1* regulation.

Further associations with MDD were found for CpG sites in the *UGT8, FNDC3B* and *SLIT2* loci. *UGT8* is a known blood biomarker gene for low mood with evidence of differential expression in human postmortem brains from mood disorder subjects ^38^. In addition, lower expression of *UGT8* have been shown in brain tissue from subjects with MDD compared with normal controls ^39^. Therefore, our study extends the current literature by highlighting that not only gene expression, but also brain-tissue based methylation in *UGT8* is linked to MDD. *FNDC3B* and *SLIT2* have been discussed in association with brain tumors ^40^, but there is no evidence for an association with neuropsychological disease.

We found stronger associations between brain-tissue based methylation levels and late-life MDD in men than in women. This finding is in line with previous studies showing sex-specific differences in serum biomarkers, mRNA expression, and brain activity of MDD cases, demonstrating that sex plays an important role in the molecular heterogeneity of MDD ^41–43^. Our findings expand the existing literature by adding DNA methylation from brain-tissue to the list of biological patterns that differ between women and men with MDD, which may have important implications for diagnosis as well as treatment strategies.

This is to our knowledge the first brain-tissue based epigenome-wide study of late-life MDD in a community-based study, and the first EWAS of MDD, which incorporates cognitive status at time of MDD diagnosis as well as at time of death. Two previous EWAS investigated the association with depressive symptoms in middle-aged and elderly people using methylation levels from whole blood ^7,8^. Beside the difference in phenotype definition, the biggest difference between these studies and ours is the tissue in which methylation was measured. In line with a previous study comparing signals from blood and brain tissue ^44^, we could not replicate the whole-blood methylation signals from ^7,8^ in our brain-tissue based EWAS (Tables S8 and S9). In two recent brain tissue-based EWAS of MDD, differential methylation was measured in 206 postmortem brain samples by enrichment-based sequencing ^9,11^. However, since in ROS/MAP methylation was measured with the Illumina HumanMethylation450 Beadchip array, loci overlapping between blood and brain in ^9^ (chr2: 208,230,169; chr9: 101,119,679; chr4: 71,632,888) were not available in ROS/MAP; therefore we could not use the dataset from Aberg et al for replication purposes. Using the same samples, Chan et al. conducted a cell type-specific EWAS, in which none of the CpGs reached epigenome-wide significance for neurons and glia or for bulk brain ^11^. Due to the different assessment of methylation (array-based versus sequencing), only two of our seven significant CpG sites were available in ^11^ and both of them were not successfully replicated.

Strengths of this study include the ROS/MAP cohort itself, which is notable for its longitudinal nature with very high follow-up rates, prospective collection of data, a community-based cohort design, and high autopsy rates. Furthermore, the twelve-month prevalence of MDD in our study population matches that in the general population^2^, which makes our findings generalizable beyond our study population. Another strength of our study is the analysis of methylation levels from brain tissue, which is the most relevant tissue for the pathophysiology of depression. In addition, we reduced the risk of confounding by cognitive status by excluding participants with a diagnosis of dementia at time of MDD assessment and by adjusting our analyses for cognitive status at the last follow-up visit (closest to methylation assessment).

The study is potentially limited by the use of bulk tissue analysis which might obscure signals from different cell populations. This problem was mitigated in our analysis by adjusting for cell-type composition. Future studies should investigate the role of *YOD1, UGT8, FNDC3B* and *SLIT2* in specific cell types from brain and investigate whether there is a causal relationship between gene dysregulation and MDD in animal models. Up to now, there is only one study analyzing cell-type specific associations between DNA methylation and MDD. However, the authors did not find any significant associations with MDD and due to the different approach of assessing differential DNA methylation (array-based versus sequencing), our most significant CpG sites were not available in their data ^11^. Therefore, there is an urgent need for more large-scale brain tissue-based EWAS of MDD to validate our and the previous ^9,11^ findings and to better understand the consequences of MDD on the human brain. Another limitation of our EWAS was the small number of MDD cases in our study population. However, to reduce the risk of false positive findings due to the imbalanced study design, we validated our findings by using different modelling approaches (limma, linear regression, permutation tests, DMR analysis).

In conclusion, we have presented evidence for brain-based DNA methylation in association with late-life MDD. We identified methylation in *YOD1, UGT8, FNDC3B* and *SLIT2* as new epigenetic factors associated with late-life MDD, which are not confounded by cognitive status or a genetic risk for MDD and stronger associated with MDD in male than in female.

## Data Availability

If you are interested in using data or specimen from the Rush Alzheimer’s Disease Center (RADC), submit a request via RADC Research Resource Sharing Hub: www.radc.rush.edu.

## Acknowledgments

The authors are grateful to the participants of the Rush Memory and Aging Project and Religious Orders Study and the Medical Research Counsel Brain Bank. Furthermore, the authors would like to thank Dr. Yiyi Ma (Columbia University Medical Center) for her valuable feedback on the manuscript.

## Competing financial interest declaration

The authors have nothing to declare.

## Web resources

Rush Alzheimer’s Disease Center Research Resource Sharing Hub: www.radc.rush.edu.

## Notes

**Funding:** AH was supported by a research fellowship from the Deutsche Forschungsgemeinschaft (DFG; HU 2731/1-1) and by the HERCULES Center (NIEHS P30ES019776). MPE was supported by NIH grant R01 GM117946. APW is supported by NIH grants R01 AG056533, VA I01 BX003853, and NIH U01 MH115484. TSW was supported by NIH grants P50 AG025688, R56 AG062256, R56 AG060757, and R01 AG056533. CR was supported by NIH grant T32 NS007480. DAB was supported by P30AG10161, R01AG15819, R01AG17917, R01AG16042, R01AG36042, U01AG61356. The funders had no role in the study design, data collection and analysis, decision to publish, or preparation of manuscript.

### Competing Interest Statement

The authors have declared no competing interest.

### Funding Statement

AH was supported by a research fellowship from the Deutsche Forschungsgemeinschaft (DFG; HU 2731/1-1) and by the HERCULES Center (NIEHS P30ES019776). MPE was supported by NIH grant R01 GM117946. APW is supported by NIH grants R01 AG056533, VA I01 BX003853, and NIH U01 MH115484. TSW was supported by NIH grants P50 AG025688, R56 AG062256, R56 AG060757, and R01 AG056533. CR was supported by NIH grant T32 NS007480. DAB was supported by P30AG10161, R01AG15819, R01AG17917, R01AG16042, R01AG36042, U01AG61356. The funders had no role in the study design, data collection and analysis, decision to publish, or preparation of manuscript.

